# Cost–Benefit Analysis of a Mobile Community-Based Electronic Medical Record System for Community Differentiated HIV Service Delivery in Lilongwe, Malawi

**DOI:** 10.64898/2025.12.10.25342019

**Authors:** Christine Kiruthu-Kamamia, Hiwot Weldemariam, Sam Truwa, Danneck Kathumba, Jacqueline Huwa, Agnes Thawani, Hannock Tweya, Wim Groot, Milena Pavlova, Caryl Feldacker

## Abstract

**Background:** Community-based differentiated service delivery (DSD) models are critical for sustaining high-quality HIV care in resource-limited settings. However, these models often rely on labor-intensive documentation processes that reduce efficiency. The Community-based ART Retention and Suppression (CARES) app, a tablet-based electronic medical record system designed through a human-centered design process, was developed to extend Malawi’s national electronic medical record system into community settings used by Lighthouse Trust’s nurse-led community ART program (NCAP). This study evaluated the cost–benefit of implementing CARES within NCAP.

**Methods:** A cost–benefit analysis was conducted from the payer perspective, using data from a time–motion study of NCAP nurses and staff observed before and after CARES implementation. Observations were collected over 28 days between 2022 and 2024. Costs included one-time development, equipment, and training expenses, and annual maintenance costs. Benefits included annual time savings for healthcare workers and reduced documentation errors. All costs were converted to USD, discounted at 5%, and modeled over a 10-year horizon. Outcomes included present value (PV), net present value (NPV), benefit–cost ratio (BCR), and break-even benefit.

**Results:** CARES reduced total time spent on observed NCAP tasks from 8 hours and 9 minutes pre-implementation to 4 hours and 23 minutes post-implementation. Annual benefits totaled $7,108, primarily from nurse time savings. The 10-year present value (PV) of benefits was $54,883, compared with PV costs of $46,499, yielding a net present value of $8,384 and a benefit–cost ratio of 1.18. CARES exceeded the annual benefit required for cost-neutrality by approximately $1,086, indicating a positive economic return under routine implementation.

**Conclusion:** CARES substantially reduced the time required for healthcare worker documentation within a routine community-based DSD model and surpassed cost-neutrality, generating a net economic benefit over 10 years. These efficiency gains create additional capacity for patient care and reinforce the platform’s operational sustainability. CARES provides a practical foundation for additional, future AI-enabled enhancements. Continued evaluation is needed to quantify clinical and system-wide benefits beyond time savings.

## INTRODUCTION

HIV remains a significant global health challenge, with sub-Saharan Africa, including Malawi, experiencing some of the highest HIV prevalence rates worldwide.(1) The World Health Organization (WHO) advocates for the adoption of differentiated service delivery (DSD) models for HIV care, which aim to provide tailored care to people living with HIV (PLHIV) regardless of their age, socioeconomic status, background, viral load levels, or ART stability.(2) Amid recent shifts in global health funding, with external aid plateauing and increasing pressure on countries to optimize limited resources, DSD models have become even more critical for sustaining access to quality HIV care while improving efficiency of service delivery.

Community-based DSD models are especially important for stable clients who can safely receive antiretroviral therapy (ART) refills and routine assessments outside overcrowded facilities.(2,3) Lighthouse Trust, Malawi’s largest ART provider, implements one such model, the nurse-led community ART program (NCAP), which provides multi-month ART refills, quarterly clinical screening, and adherence support to more than 6,000 stable clients through peer-support group sites across Lilongwe. (4) By shifting routine care into community settings, NCAP reduces travel burdens for clients and alleviates congestion at facility-based clinics.

Mobile health (mHealth), the use of mobile devices to improve healthcare delivery (5), is revolutionizing healthcare systems, including HIV management.(6–9) The WHO underscores the potential of mHealth technologies to enhance person-centered care by equipping healthcare workers with real-time, on-site data that supports more responsive, individualized decision-making and improved continuity of care.(6) In resource-constrained settings, mHealth interventions have strengthened health system efficiency by reducing documentation burdens, improving data quality, and enabling more effective task shifting.(7–10) These gains have contributed to improving HIV treatment outcomes, including ART adherence, client retention, and viral load suppression. (11,12)

In collaboration with the International Training and Education Centre for Health (I-TECH), Seattle, Washington, and Medic Mobile, Nairobi, Kenya, Lighthouse Trust developed the Community-based ART Retention and Suppression (CARES) app, a tablet-based electronic medical records system (EMRS) tailored to support NCAP. Using participatory human-centered design (HCD), the CARES app was co-created with NCAP staff, culminating in the launch of its prototype. (13–15) The app was developed to extend Malawi’s facility-based EMRS into community sites, enabling real-time point-of-care documentation, automated clinical prompts, and streamlined transfer of visit records into facility systems.

Before implementing CARES, a time-motion study was conducted to examine healthcare worker activities within NCAP. The study revealed that a significant proportion, namely 57% of the time, was spent on data entry and manual record linking. (14) These findings highlighted a clear opportunity for CARES to streamline workflows and enhance efficiency within DSD models. There is a growing body of evidence evaluating the costs and effectiveness of community-based DSD models compared with facility-based care models in low- and middle-income countries (LMICs), showing that community DSD models deliver comparable or improved clinical outcomes while reducing costs and easing the burden on health facilities.(16–19) There is also separate research assessing the costs of mHealth interventions for HIV that suggests that mHealth tools can be delivered at relatively low cost and enhance HIV care.(20–22) However, there remains a critical gap in studies that evaluate the cost-benefit of integrated mHealth tools within community-based DSD models in sub-Saharan Africa.

This gap appears wider with the emerging potential of AI-enabled mHealth and its promises to streamline service delivery, with reported early prototypes demonstrating early gains in patient monitoring efficiencies, data aggregation, and cost reductions. (23) However, for either current mHealth tools or future AI-enabled mHealth to deliver sustainable value, rigorous cost–benefit evaluation is essential, capturing both development and recurrent costs alongside productivity gains. With global health financing continuing to shrink and the urgent need for more efficient care delivery, evidence to guide policy decisions on a sustainable HIV response has become increasingly critical. To help fill this gap, we conducted a cost-benefit analysis to evaluate the economic impact of the CARES app.

## METHODS

### Study design

A cost-benefit analysis was performed from the program (payer) perspective, comparing healthcare worker costs before and after the implementation of the CARES app. We conducted a descriptive time-motion costing study observing the workload of NCAP healthcare workers (HCWs), including NCAP nurses and a data officer, as they completed NCAP-related activities and data management tasks. All nurses provide direct services to NCAP clients in the community, and the data officer is responsible for managing NCAP data, including data entry and tracking clients with missed appointments. As with many routine HCWs, some providers also performed duties outside NCAP-related tasks but were only observed during discrete NCAP tasks. We categorized observed time proportionally to estimate the share of total provider work time spent on NCAP service delivery activities that were unrelated to data management, CARES-related tasks, or EMRs to CARES data integration activities. The economic evaluation was reported in accordance with the 2022 Consolidated Health Economic Evaluation Reporting Standards (CHEERS).(24) (S1 Table).

### Setting

This study was conducted at the Lighthouse Trust Center of Excellence ART clinics in Lilongwe, Malawi. Lighthouse Trust manages five top-tier clinics across Malawi, including two main clinics in urban Lilongwe: Lighthouse (LT) and Martin Preuss Center (MPC), which together serve over 37,000 ART clients. Additionally, the Trust provides ART services at seven peri-urban satellite sites in Lilongwe District.(25–27) To ensure adherence to national ART guidelines, enhance client care and program outcomes, and streamline reporting, Lighthouse Trust utilizes the Malawi Ministry of Health’s point-of-care EMRS at all its sites. However, like other government-run clinics, each facility’s EMR operates independently, with no data sharing between clinics. NCAP services are available to clients from LT, MPC, and the seven satellite sites, but the CARES app prototype has been implemented exclusively for clients at LT and MPC.

### NCAP

Since 2016, Lighthouse Trust’s NCAP community DSD model has provided localized ART services to stable clients aged 18 and older who have been on first- or second-line treatment for at least six months with documented viral suppression.(4,13,14,28) NCAP offers monthly support group meetings, 3–6-month ART refills, and specialist referrals as needed. A client attends NCAP sites once per quarter. Previously, visit data were collected using an open data-kit (ODK)(29)-based tool and manually entered into the EMRS, a process the CARES app aims to streamline. The CARES prototype addresses several limitations of the ODK tool, including the lack of decision support, integrated care, and laboratory management, while also reducing paperwork. It enhances data transfer efficiency and improves overall client care delivery.

### CARES App

The CARES app, developed in 2022 through a user-centered design process involving clients, healthcare workers, and Ministry of Health stakeholders, was built on the open-source Community Health Toolkit (CHT)(30) to reduce costs and support scalability.(13–15) NCAP supports 6,298 clients alive in care across all participating sites, including 3,033 clients at LT and MPC sites where the CARES prototype is implemented. Designed for offline-first use in low-connectivity settings, such as NCAP and CARES, aims to extend Malawi’s national EMRS into the community. CARES supports nurses during community visits by guiding structured ART reviews with decision aids aligned to national guidelines. It enables real-time point-of-care data entry, alerts for clinical concerns, assists with medication preparation, and supports new client enrollment in the facility. Following each consultation, CARES automatically synchronizes data with the point-of-care EMR, eliminating manual data entry and maintaining efficient documentation.

### Data Collection

To facilitate time tracking for this study, Lighthouse Trust research assistants (RAs) performed activity timekeeping, observing different cadres of routine HCWs along each step of the NCAP data pathway, including NCAP file preparation, community-based data collection, data management, and manual entry into the EMRs. A data collection tool (S2 Figure) listing all activities performed during NCAP-related work enabled categorization of time and the estimation of the proportion of total provider work time spent on different NCAP tasks, including service delivery unrelated to data management, CARES-related tasks, or EMRS–CARES integration. Not all activities occur daily. RAs used the stopwatch function on their phones to record time for each task. The number of clients served was estimated for each activity, and facilities were recorded. Observations of 13 nurses and data officers were made over 17 days of NCAP service delivery between October 2022 and June 2023 for the pre-CARES period and over 11 days between March 6 and April 12, 2024, for the post-CARES period. Observations included both routine and periodic activities (S3 Table), utilizing consistent checklists and timekeeping methods. All NCAP-related activities were observed, regardless of whether CARES would impact them. Some HCWs worked across multiple facilities and were observed performing various tasks within a single day.

### Discounting

We applied a 5% annual social discount rate, consistent with recommendations for LMICs due to their typically higher rates of economic growth.(31)

### Currency, Price Date, and Conversion

All costs were recorded in Malawi Kwacha (MWK) and then converted to U.S. dollars (USD) for standardization. We applied an exchange rate of 1 USD = 1,700 MWK, consistent with the average rate during the 2023–2024 period, which aligned with the timing of the CARES implementation and post-intervention data collection.

### Costs and benefits

#### Costs

Costs were classified into four main categories: 1) Development costs referred to the one-time expenditure for application design and build, including personnel costs for app developers, and NCAP HCWs who provided feedback during development 2) Equipment costs covered the purchase of the server and tablets used for implementation 3)Training costs included the initial expenses for training HCWs on app use and 4)Annual maintenance costs were estimated at 10% of the initial development costs to account for ongoing software support, updates, and technical upkeep. In the absence of primary maintenance data, this 10% estimate reflects anticipated cost efficiencies from CARES’ open-source design and integration within the existing Lighthouse Trust IT infrastructure.

#### Benefits

The primary outcome was the annual HCW time saved (in hours/year), estimated separately for nurses and data officers. The hourly salary rate was calculated based on the HCW annual salaries. We also included reduced documentation errors as a secondary benefit attributable to CARES’ automated EMRS synchronization. Because we did not directly measure errors, we parameterized them from the literature: a 10% baseline documentation error rate(32,33), and a 50% relative reduction following EMR implementation in comparable settings. (34,35)

### Data Analysis

The time-motion study included more observations during the pre-CARES phase than during the post-CARES phase due to funding constraints, resulting in a smaller sample of observed activities post-implementation. To maintain comparability, we calculated the average time per activity by dividing the total time spent by the number of observed activities, thereby allowing consistent per-activity comparisons. We assumed that all one-time costs (development, training, and productivity loss) were incurred at the start of year 1, and that annual maintenance costs and benefits accrued at the end of each year over 10 years. We applied a 10-year analytic horizon because digital health platforms have multi-year durability, and this period allows upfront development costs to be spread over a realistic programmatic lifecycle while capturing sustained efficiency gains.(36) Present values of costs and benefits were calculated over the time horizon, including the net present value (NPV), benefit–cost ratio (BCR), and the break-even annual benefit (the minimum annual savings required for CARES to offset its costs).

### Sensitivity Analysis

One-way sensitivity analyses were conducted, varying each key parameter (development cost, training cost, productivity loss, annual maintenance, and annual benefits) within plausible ranges around their base values. Cost parameters were generally varied by ±20% from the base case, while annual maintenance was modeled at 10% to 20% of development costs, and annual benefits were adjusted based on the range of errors avoided. For each scenario, NPV and BCR were recalculated to identify the parameters exerting the most significant influence on NPV.

### Ethics

The CARES study protocol, including this costing component, was approved by the Malawi National Health Sciences Research Committee (Protocol# 21/11/2830) and the University of Washington, Seattle, USA (STUDY00013936) ethics review board. HCWs verbally consented to observations, completing routine work in routine settings. No identifiable data were collected for this costing study.

## RESULTS

Table 1 summarizes the one-time and recurring cost inputs for the CARES cost–benefit analysis. One-time costs reflect CARES development and include personnel ($21,164) covering software development, NCAP provider feedback sessions, equipment ($3,350) for the server and tablets; and initial training ($1,500) to prepare NCAP healthcare workers to use CARES. Recurring costs were limited to annual system maintenance, estimated at ($2,601) (set at 10% of development costs)).

**Table 1:** Cost categories.

| Parameter | Costs |
| --- | --- |
| **One-time Costs** |  |
| Personnel | \$21,164 |
| Equipment | \$3,350 |
| Training cost | \$1,500 |
| **Total** | **\$26,014** |
| **Recurring Costs** |  |
| Annual maintenance | \$2,601 |

Time–motion observations showed substantial reductions in the time required for NCAP tasks after CARES was introduced, with total daily task time decreasing from 8 hours and 9 minutes to 4 hours and 23 minutes (S4 Table). Table 2 summarizes the estimated annual benefits associated with implementing CARES, derived from time-motion observations and estimates of reduction in documentation errors. Time savings from the time–motion study (S4 Table) were used to calculate annual savings. CHS nurses saved 89.08 hours per year, which, at an hourly wage of $9.48, corresponded to $6,756 in annual savings for the eight CHS nurses. The data officer saved 9.67 hours per year, corresponding to minimal annual savings of $30. Improved NCAP documentation was estimated to reduce documentation errors. We estimated 12,132 NCAP visits annually for 3,033 enrolled clients and assumed a baseline documentation error rate of 10%(32,33), corresponding to approximately 1,213 errors per year. With an estimated 50% reduction(34,35) in errors following CARES implementation, an estimated 607 documentation errors were avoided each year. Using an estimated 10 minutes of data officer time per error at a wage of $3.20 per hour ($0.53 per error), this reduction translated into an additional $322 in annual savings. The combined annual monetary benefit of CARES was therefore estimated at $7,108.

**Table 2:**
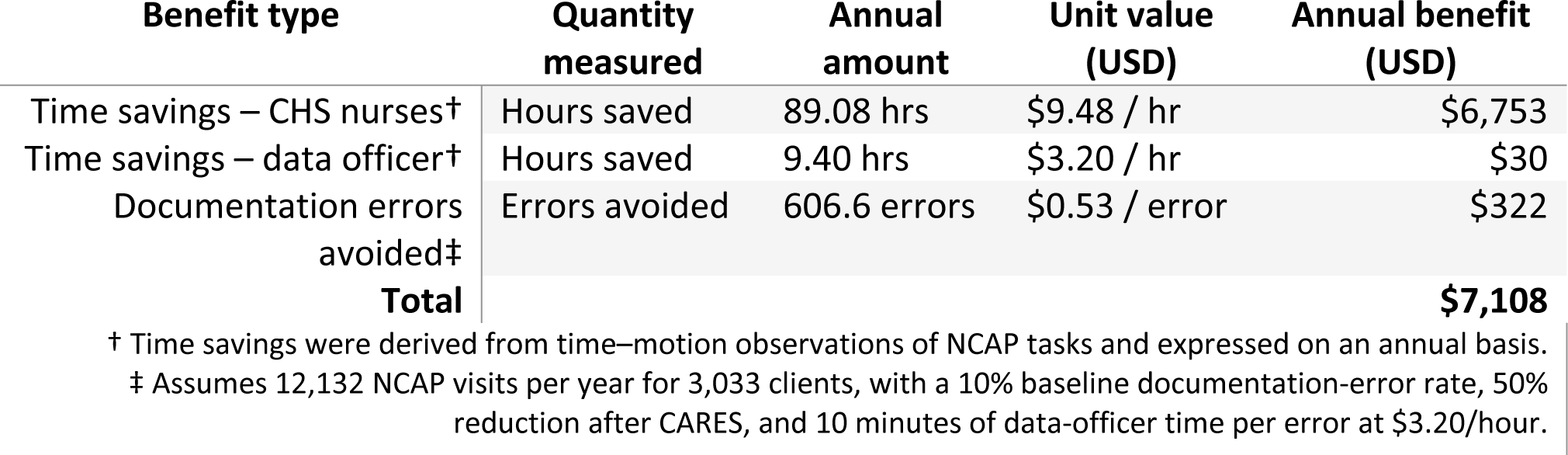
Annual Benefits.

| Benefit type | Quantity measured | Annual amount | Unit value (USD) | Annual benefit (USD) |
| --- | --- | --- | --- | --- |
| Time savings – CHS nurses† | Hours saved | 89.08 hrs | \$9.48 / hr | \$6,753 |
| Time savings – data officer† | Hours saved | 9.40 hrs | \$3.20 / hr | \$30 |
| Documentation errors avoided‡ | Errors avoided | 606.6 errors | \$0.53 / error | \$322 |
| **Total** | | | | **\$7,108** |

† Time savings were derived from time–motion observations of NCAP tasks and expressed on an annual basis.
‡ Assumes 12,132 NCAP visits per year for 3,033 clients, with a 10% baseline documentation-error rate, 50% reduction after CARES, and 10 minutes of data-officer time per error at \$3.20/hour.

Table 3 presents the cost–benefit results of CARES over a 10-year horizon at a 5% discount rate. The PV of quantified benefits was estimated at $54,883, while the combined PV of costs, including upfront development, equipment, training, and annual maintenance, totaled $46,499. This yielded an NPV of $ 8,384 and a BCR of 1,18, indicating that the discounted benefits exceeded the discounted costs. CARES also surpassed the annual benefit required to break even($6,022 per year) by approximately $1,086, demonstrating a positive economic return under base-case assumptions. One-way sensitivity analyses showed that results were most sensitive to development and annual maintenance costs, while variations in other parameters had comparatively limited impact on NPV (S5 Fig).

**Table 3.**
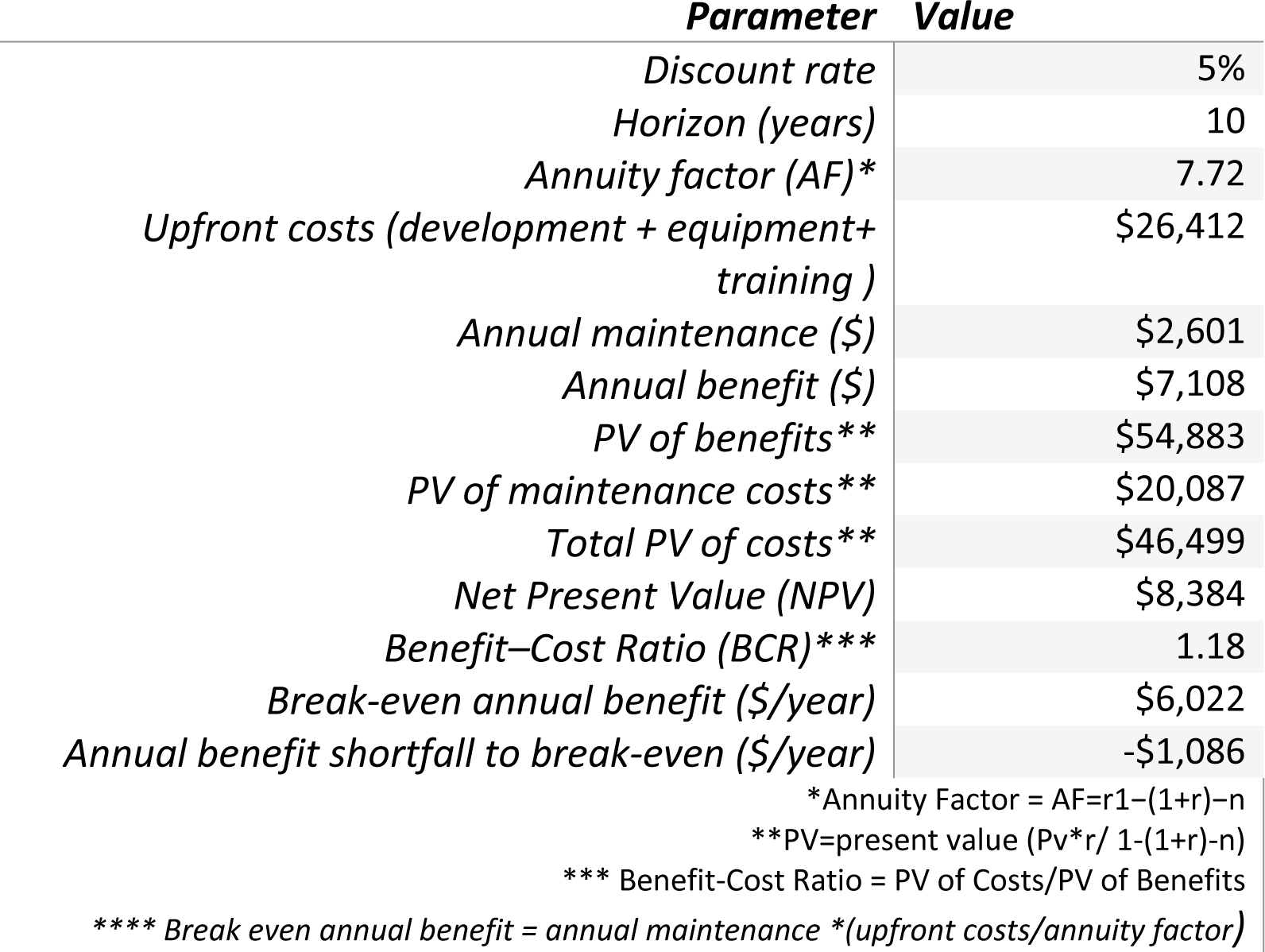
CARES Cost–Benefit Analysis Results (10-Year Base Case)

## DISCUSSION

Building on our pre-CARES deployment time-motion study(14), this analysis demonstrates that the time spent on NCAP activities and data integration decreased substantially after CARES implementation across nearly all observed tasks. Among nurses and the data officer, the total time spent on CARES-affected activities decreased by half, from approximately 8 hours pre-CARES to 4 hours post-CARES, indicating that CARES effectively reduced the time healthcare workers spent on NCAP activities including routine documentation, data integration, and reporting. From a cost–benefit perspective, this paper is novel in assessing costs from the viewpoint of a routine, large-scale DSD model implemented by a public HIV service provider in an LMIC. Over a 10-year horizon, the PV of benefits was estimated at $ 54,883, compared with PV costs of $46,499, yielding an NPV of $8,384 and a BCR of 1.18. The break-even analysis demonstrated that CARES generated more annual benefits than were required for cost-neutrality, indicating that, over a 10-year horizon, CARES is cost-beneficial, with benefits exceeding costs under routine implementation. We discuss several implications of the study and results.

First, app development and training accounted for the dominant upfront investments, while annual maintenance represented the principal recurring cost required for long-term functionality. Monetized benefits were primarily driven by nurse time savings, with smaller but meaningful contributions from data officer time and reduced documentation errors. These findings align with prior studies in LMICs, which have shown that mHealth tools streamline workflows and enhance data quality(37,38), while reaffirming that strategic investment and scale are crucial for achieving full financial sustainability. (20,22,39) CARES also streamlines MOH reporting by automatically linking NCAP data to routine EMR indicators and national M&E systems. (13,15) For health workers, CARES reduces repetitive administrative tasks, freeing time for patient care and potentially improving job satisfaction and reducing burnout. Moreover, the sensitivity analyses showed that development and maintenance costs had the greatest impact, but none of the parameter changes altered the overall direction of the findings.

Second, beyond measurable cost and time savings, CARES provides additional programmatic and clinical value that were unaccounted for in the costing exercise, but meaningfully strengthen HIV service delivery in several key ways. For clinicians, the platform is designed to enhance adherence to ART guidelines through point-of-care decision support, structured ART reviews, and clinical prompts. Given the central role of community nurses in NCAP, the workflow relief introduced by CARES may also improve job satisfaction and strengthen retention of frontline health workers, which is vital for sustaining service delivery. For data quality, CARES improves completeness, timeliness, and accuracy; reduces duplicate data entry; and facilitates drug-stock reconciliation and laboratory tracking. (15,28) For clients, it amplifies the benefits of NCAP by reducing waiting times, improving satisfaction, minimizing loss to follow-up, and enabling earlier detection of clinical problems. (15) At the program level, CARES appears to support continuity of care by ensuring that community-based services maintain the same quality standards as facilities, even in low-connectivity environments.(15) Taken together, these broader benefits strongly suggest that this cost–benefit analysis captures only one dimension of CARES’s overall value. Future CARES assessments should aim to quantify its broader health, data, and system-level impacts.

Lastly, as the mHealth field rapidly evolves, CARES provides an example of an innovation that enhances routine program delivery rather than creating a parallel or stand-alone system, providing a solid foundation from which to continue digital innovation. Future enhancements and new technologies could aid efficiency gains by leveraging AI-enabled capabilities, such as client record matching and deduplication, optical character recognition (OCR) of legacy paper forms, automated anomaly detection, and predictive analytics, to identify clients at risk of attrition, thereby extending efficiency gains and strengthening service performance. (40,41) At the same time, AI integration brings new dimensions of governance and regulatory requirements, including model licensing, data oversight, and performance monitoring(41)-likely adding development costs to future iterations. Developing a robust evidence base around the actual cost–benefit and equity implications of AI adoption in resource-limited settings will be key to guiding sustainable scale-up.

### Limitations

This study has several limitations, largely due to its implementation in a routine setting and limited funding. NCAP personnel performed both NCAP and non-NCAP duties, and individual activities could not always be isolated, introducing imprecision in attributing time specifically to CARES. The observation period, particularly post-CARES, was limited, and not all clerks were observed; therefore, some activities and efficiency changes may not have been captured. Furthermore, observations also occurred during shifts in staffing and skill mix, which may have influenced measured differences between periods. Because many tasks were performed by experienced staff, the findings may not be generalizable to settings with less experienced or lower-cadre personnel. Estimates of reductions in documentation errors relied on literature-based assumptions rather than empirical measurement, thus limiting the precision of the benefit calculations. Additionally, CARES did not fully synchronize with the EMR during the evaluation period, requiring manual data aggregation, which likely reduced the observed time savings. Despite these limitations, the triangulation of time-motion data, program cost information, and sensitivity analyses provides a credible approximation of CARES’ efficiency gains.

## Conclusion

This study evaluated whether integrating an mHealth tool into a routine community-based DSD model could improve efficiency, data quality, and continuity of care. Our analysis shows that CARES not only achieved but exceeded cost-neutrality, generating a net economic benefit over the 10-year horizon. The time savings generated by CARES translate into additional workforce capacity that can be redirected toward higher-value patient care activities. Within NCAP, where registered community nurses play a central role in clinical assessment, counseling, and care coordination, this added capacity can enhance client follow-up, facilitate tracing of individuals lost to care, and strengthen continuous quality improvement efforts.

When accumulated at scale, these operational gains may also support the retention and well-being of frontline health workers, contributing to a more stable workforce and sustained equitable service coverage. Overall, CARES demonstrates how well-designed digital tools can augment, not replace, human capacity within differentiated HIV service delivery models. By strengthening routine operations, the platform provides a practical and scalable foundation for future digital innovations, including AI-enabled decision-support tools, as HIV programs continue to navigate constrained resources and evolving care needs.

## Data Availability

All data underlying the results presented in this manuscript will be made available.

## SUPPORTING INFORMATION

**S1 Table CHEERS 2022 Checklist**

**S2 Fig CARES time motion study observation tool**

**S3 Table NCAP Activities Observed**

**S4 Table Average times pent on activities pre-and post CARES**

**S5 Figure Tornado diagram of the sensitivity analysis**

## ACKNOWLEDGMENTS

The authors would like to thank the Lighthouse Trust community health services program, and the research department for their partnership in implementing the CARES application.

## REFERENCES

1. UNAIDS. The urgency of now: AIDS at a crossroads. Geneva: Joint United Nations Programme on HIV/AIDS [Internet]. 2024. Available from: https://www.unaids.org/sites/default/files/media_asset/2024-unaids-global-aids-update_en.pdf

2. World Health Organization. WHO guideline on HIV service delivery: updated guidance on the integration of diabetes, hypertension and mental health services, and interventions to support adherence to antiretroviral therapy. 2025; Available from: https://iris.who.int/server/api/core/bitstreams/f6e902a0-b37b-4340-8296-39d91f3321bb/content

3. Hagey JM, Li X, Barr-Walker J, Penner J, Kadima J, Oyaro P, et al. Differentiated HIV care in sub-Saharan Africa: a scoping review to inform antiretroviral therapy provision for stable HIV-infected individuals in Kenya. AIDS Care [Internet]. 2018 Dec 2 [cited 2021 Mar 27];30(12):1477–87. Available from: 10.1080/09540121.2018.1500995

4. Sande O, Burtscher D, Kathumba D, Tweya H, Phiri S, Gugsa S. Patient and nurse perspectives of a nurse-led community-based model of HIV care delivery in Malawi: a qualitative study. BMC Public Health [Internet]. 2020 May 14;20(1):685. Available from: 10.1186/s12889-020-08721-6

5. WHO Global Observatory for eHealth. mHealth: new horizons for health through mobile technologies: second global survey on eHealth. 2011; Available from: https://apps.who.int/iris/handle/10665/44607

6. World Health Organization, World Bank Group, OECD. Delivering Quality Health Services: A Global Imperative for Universal Health Coverage [Internet]. Geneva: World Health Organization; 2018 [cited 2024 Apr 7]. Available from: http://hdl.handle.net/10986/29970

7. Odendaal WA, Anstey Watkins J, Leon N, Goudge J, Griffiths F, Tomlinson M, et al. Health workers’ perceptions and experiences of using mHealth technologies to deliver primary healthcare services: a qualitative evidence synthesis. Cochrane Database Syst Rev [Internet]. 2020 Mar 26 [cited 2024 Mar 3];2020(3):CD011942. Available from: https://www.ncbi.nlm.nih.gov/pmc/articles/PMC7098082/

8. Burnett SM, Wun J, Evance I, Davis KM, Smith G, Lussiana C, et al. Introduction and Evaluation of an Electronic Tool for Improved Data Quality and Data Use during Malaria Case Management Supportive Supervision. Am J Trop Med Hyg [Internet]. 2019 Apr 1 [cited 2024 Apr 7];100(4):889–98. Available from: https://europepmc.org/articles/PMC6447118

9. Waters E, Rafter J, Douglas GP, Bwanali M, Jazayeri D, Fraser HSF. Experience Implementing a Point-of-Care Electronic Medical Record System for Primary Care in Malawi. In: MEDINFO 2010 [Internet]. IOS Press; 2010 [cited 2024 Mar 10]. p. 96–100. Available from: https://ebooks.iospress.nl/doi/10.3233/978-1-60750-588-4-96

10. World Health Organization. WHO guideline: recommendations on digital interventions for health system strengthening [Internet]. Geneva: World Health Organization; 2019 [cited 2024 Apr 1]. Available from: https://iris.who.int/handle/10665/311941

11. Lalla-Edward ST, Mashabane N, Lynsey Steward-Isherwood, Scott L, Scott L, Kyle Fyvie, et al. Technical Feasibility and Acceptability of the iThemba Life mobile health application to support engagement in HIV care and viral load suppression: A pilot study in Johannesburg, South Africa (Preprint). JMIR Form Res. 2020 Nov 25;

12. Demena BA, Artavia-Mora L, Ouedraogo D, Thiombiano BA, Wagner N. A Systematic Review of Mobile Phone Interventions (SMS/IVR/Calls) to Improve Adherence and Retention to Antiretroviral Treatment in Low-and Middle-Income Countries. AIDS Patient Care STDs [Internet]. 2020 Feb [cited 2023 Sept 11];34(2):59–71. Available from: https://www.liebertpub.com/doi/10.1089/apc.2019.0181

13. Feldacker C, Mugwanya R, Irongo D, Kathumba D, Chiwoko J, Kitsao E, et al. A Community-Based, Mobile Electronic Medical Record System App for High-Quality, Integrated Antiretroviral Therapy in Lilongwe, Malawi: Design Process and Pilot Implementation. JMIR Form Res [Internet]. 2023 Nov 10 [cited 2025 Oct 7];7:e48671. Available from: https://pmc.ncbi.nlm.nih.gov/articles/PMC10674144/

14. Feldacker C, Usiri J, Kiruthu-Kamamia C, Waehrer G, Weldemariam H, Huwa J, et al. Crossing the digital divide: the workload of manual data entry and integration between mobile health applications and eHealth infrastructure. Oxf Open Digit Health [Internet]. 2024 Nov 28 [cited 2025 Jan 26];2(Supplement_2):ii9–17. Available from: 10.1093/oodh/oqae025

15. Kiruthu-Kamamia C, Berner-Rodoreda A, O’Bryan G, Sande O, Huwa J, Thawani A, et al. Acceptability and feasibility of a mobile electronic medical record system for community-based antiretroviral therapy in Lilongwe, Malawi: A rapid qualitative analysis. PLOS ONE [Internet]. 2025 May 23 [cited 2025 Dec 8];20(5):e0303416. Available from: https://journals.plos.org/plosone/article?id=10.1371/journal.pone.0303416

16. Liang A, Wilson-Barthes M, Galárraga O. Cost-effectiveness of differentiated care models that incorporate economic strengthening for HIV antiretroviral therapy adherence: a systematic review. Cost Eff Resour Alloc [Internet]. 2024 May 24 [cited 2025 June 25];22(1):46. Available from: 10.1186/s12962-024-00557-w

17. Guthrie T, Muheki C, Rosen S, Kanoowe S, Lagony S, Greener R, et al. Similar costs and outcomes for differentiated service delivery models for HIV treatment in Uganda. BMC Health Serv Res [Internet]. 2022 Nov 3 [cited 2025 June 25];22(1):1315. Available from: 10.1186/s12913-022-08629-4

18. Roberts DA, Tan N, Limaye N, Irungu E, Barnabas RV. Cost of Differentiated HIV Antiretroviral Therapy Delivery Strategies in Sub-Saharan Africa: A Systematic Review. JAIDS J Acquir Immune Defic Syndr [Internet]. 2019 Dec [cited 2025 June 25];82:S339. Available from: https://journals.lww.com/jaids/fulltext/2019/12003/Cost_of_Differentiated_HIV_Antiretroviral_Therapy.23.aspx

19. Larson BA, Pascoe SJ, Huber A, Long LC, Murphy J, Miot J, et al. Will differentiated care for stable HIV patients reduce healthcare systems costs? J Int AIDS Soc [Internet]. 2020 July 20 [cited 2021 Mar 27];23(7). Available from: https://www.ncbi.nlm.nih.gov/pmc/articles/PMC7370539/

20. Iribarren SJ, Cato K, Falzon L, Stone PW. What is the economic evidence for mHealth? A systematic review of economic evaluations of mHealth solutions. PLoS ONE [Internet]. 2017 Feb 2 [cited 2022 Oct 21];12(2):e0170581. Available from: https://www.ncbi.nlm.nih.gov/pmc/articles/PMC5289471/

21. Chen Y, Ronen K, Matemo D, Unger JA, Kinuthia J, John-Stewart G, et al. An Interactive Text Messaging Intervention to Improve Adherence to Option B+ Prevention of Mother-to-Child HIV Transmission in Kenya: Cost Analysis. JMIR MHealth UHealth [Internet]. 2020 Oct 2 [cited 2022 Oct 21];8(10):e18351. Available from: https://www.ncbi.nlm.nih.gov/pmc/articles/PMC7568211/

22. Babigumira JB, Barnhart S, Mendelsohn JM, Murenje V, Tshimanga M, Mauhy C, et al. Cost-effectiveness analysis of two-way texting for post-operative follow-up in Zimbabwe’s voluntary medical male circumcision program. PloS One. 2020;15(9):e0239915.

23. Esteva A, Robicquet A, Ramsundar B, Kuleshov V, DePristo M, Chou K, et al. A guide to deep learning in healthcare. Nat Med [Internet]. 2019 Jan [cited 2025 Dec 8];25(1):24–9. Available from: https://www.nature.com/articles/s41591-018-0316-z

24. Husereau D, Drummond M, Augustovski F, Bekker-Grob E de, Briggs AH, Carswell C, et al. Consolidated Health Economic Evaluation Reporting Standards 2022 (CHEERS 2022) statement: updated reporting guidance for health economic evaluations. BMJ [Internet]. 2022 Jan 11 [cited 2022 Nov 23];376:e067975. Available from: https://www.bmj.com/content/376/bmj-2021-067975

25. World Health Organization. The Lighthouse: a centre for comprehensive HIV/AIDS treatment and care in Malawi: case study [Internet]. World Health Organization; 2004 [cited 2023 Sept 11]. Available from: https://apps.who.int/iris/handle/10665/43011

26. Phiri S, Neuhann F, Glaser N, Gass T, Chaweza T, Tweya H, et al. The path from a volunteer initiative to an established institution: evaluating 15 years of the development and contribution of the Lighthouse trust to the Malawian HIV response. BMC Health Serv Res. 2017 Aug 9;17(1):548.

27. Lighthouse Trust. Centres of Excellence [Internet]. [cited 2023 Sept 11]. Available from: https://www.mwlighthouse.org/about-lighthouse-trust/centres-of-excellence

28. Orii L, Wilson KS, Huwa J, Kiruthu-Kamamia C, Sande O, Thawani A, et al. “They gave us the right to choose.” A qualitative study of preferences for differentiated service delivery location among recipients of antiretroviral therapy at Lighthouse Trust in Lilongwe Malawi. PLOS ONE [Internet]. 2025 Feb 6 [cited 2025 Sept 29];20(2):e0296531. Available from: https://pmc.ncbi.nlm.nih.gov/articles/PMC11801716/

29. ODK - Collect data anywhere [Internet]. [cited 2025 Dec 8]. Available from: https://getodk.org

30. Community Health Toolkit [Internet]. 2025 [cited 2025 Dec 8]. Why the Community Health Toolkit? Available from: https://docs.communityhealthtoolkit.org/why-the-cht/

31. Haacker M, Hallett TB, Atun R. On discount rates for economic evaluations in global health. Health Policy Plan. 2020 Feb 1;35(1):107–14.

32. Msiska KEM, Kumitawa A, Kumwenda B. Factors affecting the utilisation of electronic medical records system in Malawian central hospitals. Malawi Med J [Internet]. 2017 [cited 2025 Sept 22];29(3):247–53. Available from: https://www.ajol.info/index.php/mmj/article/view/163200

33. Castelnuovo B, Kiragga A, Afayo V, Ncube M, Orama R, Magero S, et al. Implementation of Provider-Based Electronic Medical Records and Improvement of the Quality of Data in a Large HIV Program in Sub-Saharan Africa. PLOS ONE [Internet]. 2012 Dec 17 [cited 2025 Sept 22];7(12):e51631. Available from: https://journals.plos.org/plosone/article?id=10.1371/journal.pone.0051631

34. Uslu A, Stausberg J. Value of the Electronic Medical Record for Hospital Care: A Review of the Literature. J Healthc Eng [Internet]. 2011 [cited 2025 Sept 22];2(3):639549. Available from: https://onlinelibrary.wiley.com/doi/abs/10.1260/2040-2295.2.3.271

35. Naamneh R, Bodas M. The effect of electronic medical records on medication errors, workload, and medical information availability among qualified nurses in Israel– a cross sectional study. BMC Nurs [Internet]. 2024 Apr 24 [cited 2025 Sept 22];23(1):270. Available from: 10.1186/s12912-024-01936-7

36. Bertram MY, Lauer JA, Stenberg K, Edejer TTT. Methods for the Economic Evaluation of Health Care Interventions for Priority Setting in the Health System: An Update From WHO CHOICE. Int J Health Policy Manag [Internet]. 2021 Jan 20 [cited 2025 Nov 26];10(11):673–7. Available from: https://pmc.ncbi.nlm.nih.gov/articles/PMC9278384/

37. Boyce SP, Nyangara F, Kamunyori J. A mixed-methods quasi-experimental evaluation of a mobile health application and quality of care in the integrated community case management program in Malawi. J Glob Health [Internet]. [cited 2024 Mar 13];9(1):010811. Available from: https://www.ncbi.nlm.nih.gov/pmc/articles/PMC6594718/

38. Ødegård ES, Langbråten LS, Lundh A, Linde DS. Two-way text message interventions and healthcare outcomes in Africa: Systematic review of randomized trials with meta-analyses on appointment attendance and medicine adherence. PloS One. 2022;17(4):e0266717.

39. Shiferaw MB, Endalamaw D, Hussien M, Agegne M, Amare D, Estifanos F, et al. Viral suppression rate among children tested for HIV viral load at the Amhara Public Health Institute, Bahir Dar, Ethiopia. BMC Infect Dis. 2019 May 14;19(1):419.

40. Khan EA. Lessons from a seven-year experience of paediatric HIV in Pakistan: A single centre experience. JPMA J Pak Med Assoc. 2017 Jan;67(1):105–10.

41. Kamitani E, Koenig LJ, Sullivan P. Transformative potential of artificial intelligence in US CDC HIV interventions: balancing innovation with health privacy. AIDS Lond Engl. 2025 Aug 1;39(10):1311–21.

